# Enrichment of Repeat Expansions in *FGF14* Associated with Amyotrophic Lateral Sclerosis

**DOI:** 10.64898/2026.08.16.26351538

**Authors:** Sherie Ma, Phillip K. West, Anne Trinh, Alicia Yang, Egor Dolzhenko, Ahmad Al Kheifat, Aminah Ali, Alfredo Iacoangeli, Ted Wong, P. Anthony Akkari, Nicole Ellis-Ovadia, Mohd Faruq, Ammar Al-Chalabi, Matthew B. Harms, Terry D. Heiman-Patterson, Richard Bedlack, Masha Stromme

## Abstract

Amyotrophic Lateral Sclerosis (ALS) is a neurodegenerative disease characterised by progressive motor neuron loss and corticospinal tract degeneration. The genetic landscape of ALS is complex, with increasing recognition of shared genetic and phenotypic features with other neurodegenerative conditions, particularly those involving repeat expansions. Given that repeat expansions in disorders like spinocerebellar ataxia type 27B (SCA27B), caused by an intronic GAA repeat expansion in Fibroblast Growth Factor 14 (*FGF14*), are recognised to extend beyond cerebellar ataxia with frequent pyramidal signs, we hypothesised that *FGF14* repeat expansions might also contribute to ALS and degeneration of corticospinal pathways, and sought to investigate whether repeat length is associated with clinical phenotype. We screened 62 individuals with ALS using PacBio HiFi long-read whole-genome sequencing and compared repeat-size distributions with 256 healthy controls from the Human Pangenome Reference Consortium. Repeat expansions were confirmed using flanking PCR and repeat-primed PCR. We identified pathogenic-range *FGF14* GAA ≥250 expansions, the established threshold for SCA27B, in 3/62 ALS cases (4.8%) and none in controls. Further analysis revealed that GAA expansions ≥200 repeats were enriched in ALS compared to controls (8.1% vs 0.4%; *p* = 0.0013), suggesting a broader pathogenic spectrum for *FGF14* GAA repeats in ALS. In contrast, GAAGGA expansions were not significantly associated. Expanded pure GAA alleles were predicted to form triplex (H-DNA) structures, with the repeat-containing isoform (1B) being the predominant *FGF14* transcript in motor neurons. These findings demonstrate that *FGF14* GAA repeat expansions extend into the motor neuron disease spectrum.

## Introduction

Amyotrophic Lateral Sclerosis (ALS) is a progressive neurodegenerative disorder affecting motor neurons in the brain and spinal cord, leading to muscle atrophy, paralysis, and death, typically within 2–5 years of symptom onset (1). Approximately 10–15% of ALS cases have a family history of the disease, with the remainder being sporadic. Even in apparently sporadic disease, twin and population-based studies indicate a heritability of around 60% (2, 3). 24 genes have been identified as definitely causative for ALS (Clinical Genome Resource, https://search.clinicalgenome.org/kb/affiliate/10096, accessed Feb 2026) including *C9ORF72, SOD1, TARDBP*, and *FUS* representing the most common genetic causes (4).

Repeat expansion disorders represent a major class of genetic diseases, many of which lead to neurodegenerative or neurodevelopmental conditions. These disorders are characterised by the pathological expansion of short tandem DNA repeats, which can lead to toxic RNA gain-of-function, protein aggregation, repeat associated non-AUG (RAN) translation of toxic proteins, or loss-of-function due to transcriptional silencing or altered splicing (5, 6). The identification of repeat expansions, such as the GGGGCC hexanucleotide repeat in *C9ORF72*, has advanced our understanding of the genetics of ALS pathogenesis (7, 8). Long-read whole-genome sequencing is capable of detecting these complex genomic variations, which are often missed by conventional short-read sequencing due to their repetitive nature and size (9, 10).

One such example is spinocerebellar ataxia type 27B (SCA27B), caused by an intronic GAA repeat expansion in the Fibroblast Growth Factor 14 (*FGF14*) gene. Although primarily characterised by cerebellar ataxia, recent clinical observations indicate that SCA27B patients frequently present with pyramidal signs, a hallmark of corticospinal tract involvement (11). Given that degeneration of the corticospinal pathways is a defining pathological feature of ALS, this clinical bridge between SCA27B and motor neuron disease (MND) provides a rationale to investigate whether *FGF14* repeat expansions also contribute to ALS. We hypothesised that *FGF14* repeat expansions might be associated with ALS. *FGF14* encodes a protein that plays a crucial role in neuronal excitability and synaptic plasticity, particularly in the cerebellum (12, 13). Recently, it was demonstrated that expansions of GAA repeats are pathogenic for a late-onset form of SCA27, termed SCA27B (14, 15). The precise diagnostic threshold of GAA repeat sizes is still under investigation. While expansion tracts larger than 250 repeats are considered pathogenic for SCA27B (16), repeats between 200-249 units may also contribute to disease (17–19) have recently been associated with multiple system atrophy (19, 20).

In this study, we performed long-read whole-genome sequencing in a cohort of ALS participants. We report the enrichment of GAA and GAAGGA repeat expansions within the *FGF14* gene in a subset of this cohort. RNA-seq analysis demonstrated an increased expression of *FGF14* in the motor cortex of a separate ALS dataset. We characterised the potential pathogenic mechanisms of the GAA expansion through *in silico* analyses of non-B DNA structures and investigated the expression patterns of *FGF14* isoforms in motor neurons. Our findings provide evidence for a novel genetic aetiology in ALS, suggesting a parallel with other repeat expansion disorders and highlighting the importance of long-read sequencing in uncovering complex genetic variations in neurodegenerative diseases.

## Materials and Methods

### ALS Cohort and Sample Preparation

Written informed consent was obtained from each participant for publication of anonymised information in this study. Participants in this ALS cohort were from USA (80.6%), Australia (16.1%), and India (3.2%). Almost all participants (96.7%; 60/62) did not have a known family history of ALS and were considered to have sporadic disease. Comparison to healthy control *FGF14* repeat sizes was performed using alleles called for the *FGF14* locus from PacBio HiFi long-read sequencing of 256 healthy controls from the Human Pangenome Reference Consortium (HPRC).

### Long-Read Whole-Genome Sequencing

Long read genome sequencing was undertaken by the following providers, using standard operating procedures as recommended by the manufacturer (www.pacb.com): Sequencing and Genomics Technologies Core Facility (Duke University), Genomic Technologies Group (Garvan Institute of Medical Research), Next Level Genomics Pty Ltd (Singapore), and Australian Genome Research Facility. Whole blood samples were prepared for high molecular weight DNA extraction and library preparation (https://www.pacb.com/wp-content/uploads/Procedure-checklist-Extracting-HMW-DNA-from-human-whole-blood-using-Nanobind-kits.pdf). Genomic DNA quality and concentration were assessed using the Tapestation 2200 Nucleic Analyser for sample quality and Veriskan to measure concentration, FEMTO Pulse (Agilent Technologies) and Qubit dsDNA HS reagents Assay kit (Thermo Fisher Scientific), according to manufacturer standards and protocols. Sequencing was performed on the PacBio Revio HiFi system with the Revio Polymerase Kit, following the Revio SMRT Link setup.

The PacBio HiFi-human-WGS-WDL workflow was used to analyse tandem repeats. Briefly, genomic reads were aligned to the reference genome using pbmm2 (1.10.0), and tandem repeat analysis was performed using TRGT (0.9.0). Quality control was maintained by validating sequence length distributions and ensuring a minimum depth of coverage of 20X across the target regions and minimum base quality of 20. An optimised version of the PacBio HiFi-human-WGS-WDL workflow was implemented in the proprietary Deep Integrated Genomics Analysis Platform (DiGAP™) pipeline (GenieUs Genomics).

For one ALS case (GAA-1, Table 1), associated short-read whole-genome sequencing data (Illumina Novaseq 6000 S4 PE150) were aligned to the hg38 reference genome and processed following the GATK (v4.5.0.0) Best Practices pipeline, including bwa-mem for alignment, duplicate marking and base quality score recalibration. Short tandem repeats were then genotyped using ExpansionHunter (v5.0.0).

**Table 1.** Summary of the detection and confirmation of *FGF14* repeat expansions in five expansion carriers and one non-carrier by long-read sequencing and repeat-primed PCR.

| Carrier ID | DiGAP™ long-read motif counts | UCG repeat-primed and cross-repeat PCR | ExpansionHunter short-read |
| --- | --- | --- | --- |
| GAA-1 | 319, 12 GAA<br>0, 0 GAAGGA | 325, 11 GGA | 79, 14 GAA |
| GAA-2 | 262, 12 GAA<br>0, 0 GAAGGA | 272, 11 GGA | - |
| GAA-3 | 385, 43 GAA<br>0, 0 GAAGGA | 387, 36 GGA | - |
| GAA-4 | 213, 10 GAA<br>0, 0 GAAGGA | - | - |
| GAA-5 | 202, 81 GAA<br>0, 0 GAAGGA | - | - |
| GAAGGA-1 | 21, 12 GAA<br>143, 0 GAAGGA | 326, 11 (non pure-GAA) | - |
| GAAGGA-2 | 16, 10 GAA<br>144, 0 GAAGGA | 323, 9 (non pure-GAA) | - |
| NEG-1 | 23, 9 GAA<br>0, 0 GAAGGA | 22, 8 GAA | - |

### Association Testing for Repeat Expansions

Carrier frequencies for GAA and GAAGGA repeat expansions were compared between ALS cases and healthy controls using 2×2 contingency tables at repeat-length thresholds binned in 50-repeat increments. To test the hypothesis that large expansions are enriched in ALS, statistical association was assessed using one-sided Fisher’s exact tests. Odds ratios were calculated from the contingency tables, and 95% confidence intervals were estimated using the Woolf log-odds method. For thresholds with zero control carriers, a Haldane–Anscombe correction was applied before odds ratio and confidence interval estimation.

### Clinical Repeat Expansion Analysis by Repeat-Primed PCR and Cross-Repeat PCR

*FGF14* (SCA27B) repeat expansion analysis was undertaken by The University of Chicago Genetic Services Laboratory (for cases GAA-1, GAA-2, GAA-3, GAAGGA-1, GAAGGA-2, and NEG-1 (a participant with ALS that was not a carrier of an *FGF14* expansion, included as a negative control)). Repeat sizing for the *FGF14* gene was performed by standard flanking-PCR (F-PCR) and capillary electrophoresis followed by repeat primed-PCR (RP-PCR) and capillary electrophoresis for cases with ≥200 repeats. F-PCR was used to amplify across the repeat region and RP-PCR was used to amplify within the repeat region. RP-PCR was performed using a fluorescently labelled primer specific to the target of interest, a “repeat primer” consisting of multiple repeats in tandem, and an anchor primer specific to a tail attached to the repeat primer (21). The presence of a ‘ladder’ of repeat size products was used to determine the GAA repeat nature. Sizing of the repeats was performed using the F-PCR products sized by either capillary electrophoresis or by the Agilent Tapestation. Expansions larger than 380 repeats can be detected but may not be sized by this assay. For repeat sizes 40 and above the accuracy of the assay is +/− 10 repeats and for repeat sizes less than 40 the accuracy of the assay is +/− one repeat.

### *In Silico* Analysis of Repeat Expansions

#### Allele-specific sequence construction

For all *in silico* analyses, allele-specific DNA sequences were derived from long-read sequencing data to preserve true repeat composition and length. For each allele, a representative repeat-spanning HiFi read was identified from TRGT-aligned BAM files filtered to the chr13 locus. Repeat expansion boundaries were determined directly from the read sequence. Because expanded alleles vary in length and disrupt reference coordinate correspondence downstream of the repeat, reference genomic coordinates alone could not be used to define flanking regions. Instead, sequence context was defined relative to the read-derived expansion boundaries. Each allele-specific sequence was extended by ±1000 bp upstream and downstream of the expansion, yielding a single contiguous FASTA sequence per allele for downstream analysis.

#### R-loop formation analysis

R-loop–forming sequence (RLFS) prediction was performed using QmRLFS-finder (http://r-loop.org/?pg=qmrlfs-finder; accessed July 2025), a sequence-based tool that identifies genomic regions with features associated with R-loop formation. Allele-specific FASTA sequences generated as described above were submitted to the QmRLFS-finder web server using default parameters and prediction models (m1 and m2; quick mode). Predicted RLFS calls were summarised for each allele based on the presence or absence of RLFS motifs within the input sequence. This sequence-anchored approach was used to preserve allele-specific sequence context and avoid inaccuracies introduced by reference-based coordinate extraction in expanded repeat regions.

#### H-DNA (triplex) formation analysis

To assess the potential for intramolecular triplex DNA (H-DNA) formation within repeat-expanded alleles, we performed computational prediction of non-B DNA motifs using the non-B DNA Motif Search Tool (nBMST) web server (https://nonb-abcc.ncifcrf.gov/apps/nBMST/; accessed July 2025). nBMST identifies sequence motifs associated with alternative DNA conformations, including mirror repeats capable of forming intramolecular triplex structures. Allele-specific FASTA sequences were analysed with detection of mirror repeats and triplex-forming motifs enabled. For each allele, predicted mirror repeat regions were extracted along with their coordinates relative to the input sequence, strand orientation, and motif length. Overlapping mirror repeats were retained to capture continuous or nested triplex-forming potential across expanded repeat tracts. Other non-B DNA motif classes detected by nBMST were not considered in this analysis.

### *FGF14* Isoform Expression Analysis

#### RNA-seq data acquisition and isoform quantification

##### AnswerALS dataset

A cohort of 95 RNA-seq samples (28 control, 67 ALS) from iPSC-derived spinal motor neurons was obtained from the AnswerALS consortium (22). These samples had been previously prepared using the TruSeq Stranded Total RNA kit, sequenced on the Illumina NovaSeq 6000 platform. RNAseq data was aligned to the GRCh38 reference genome using Hisat2 (v.2.2.1). Exon-level quantification was performed using featureCounts (Subread v2.0.2) guided by a custom UCSC RefSeq annotation (GRCh38) of the *FGF14* for both the canonical (isoform 1A; NM_004115) and alternate transcript (isoform 1B; NM_175929). Reads were assigned to an exon based on a minimum overlap of 20 bp and final counts were generated using a union-based overlap model to account for the stranded nature of the original library preparation. Exon-level counts were normalised by the exon length and scaled to reflect relative isoform usage, and the normalised counts for isoform 1A and isoform 1B were compared within each sample using a paired Wilcoxon signed-rank test. For differential gene expression of *FGF14* in ALS versus healthy controls, scaled TPMs for FGF14 were examined for the AnswerALS cohort (174 control, 458 ALS) using the AnswerALS Visualization Tool (22).

##### King’s College London dataset

Post-mortem motor cortex tissue was obtained from the MRC London Neurodegenerative Diseases Brain Bank at the Institute of Psychiatry, Psychology & Neuroscience, King’s College London (23). Ethical approval was granted by the local research ethics committee at King’s College London and the MRC London Neurodegenerative Diseases Brain Bank. Bulk RNA-seq analysis of human motor cortex tissue (n=101; n=25 controls and n=76 ALS) was performed. Tissue was flash-frozen and stored at −80°C. From each case, 100 mg of tissue was excised, and total RNA was isolated from the same tissue block for bulk RNA sequencing (Supplementary Table 1). Bulk RNA sequencing was performed on total RNA extracted from post-mortem human motor cortex tissue. Reads were quality-controlled, aligned to the human reference genome (GRCh38), and summarised at the gene level. Differential expression analysis comparing ALS cases with controls was conducted with adjustment for age, sex, post-mortem delay, and RNA integrity.

## Results

### Identification and confirmation of *FGF14* repeat expansions in participants with ALS

Long-read whole-genome sequencing identified heterozygous GAA and GAAGGA repeat expansions in *FGF14* in a cohort of 62 ALS participants (Table 1, Fig. 1), located in the hg38 reference genome at chr13:102,161,576-102,161,726, an intronic region associated with the alternate *FGF14* isoform 1B. Consistent with the established pathogenic range for SCA27B (16, 24), GAA expansions of ≥250 repeats were found in 3 of 62 (4.8%) ALS cases (Table 1). These pathogenic-range expansions were absent in our cohort of 256 healthy controls (Human Pangenome Reference Consortium, HPRC; Supplementary Fig. 2). In addition, we also identified 2 ALS cases with GAA repeat expansions between 200-250 repeats, and 2 ALS cases with expanded GAAGGA repeats (Table 1, Fig. 1).

**Figure 1.**
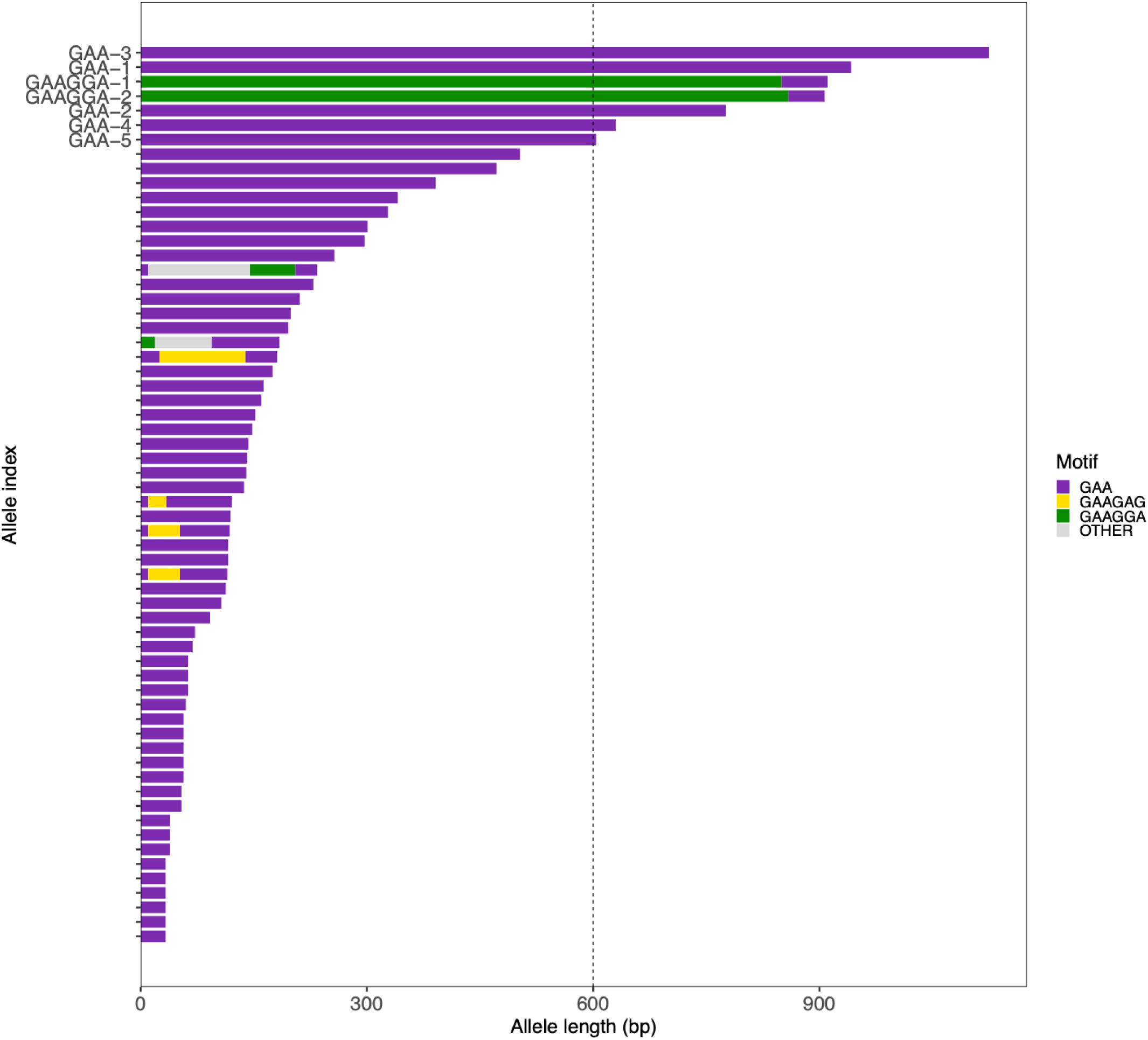
Characterization of the size and motif distribution of the *FGF14* repeat expansion in a cohort of 62 individuals with ALS. The dashed line indicates an allele length of 600 bp (200 GAA repeats).

Repeat expansion size in *FGF14* was confirmed in 6 ALS cases (three GAA expansion carriers with >250 repeats, two GAAGGA expansion carriers, and one non-carrier as a negative control) using repeat-primed PCR, and the reported size was similar to that detected by long-read sequencing for GAA and GAAGGA motifs (Table 1). For one sample, we also assessed concurrent short-read whole-genome sequencing with ExpansionHunter, which significantly under detected the number of GAA repeats in the *FGF14* gene (79 versus 319 repeats).

### Repeat expansions of GAA in *FGF14* are associated with ALS

To investigate the spectrum of *FGF14* GAA repeat involvement in ALS, we analysed repeat-size distributions across both ALS participants and healthy controls (Fig. 2A). We found a significant enrichment of GAA expansions ≥200 repeats in the ALS cohort (8.1%, 5/62 cases) compared to healthy controls (0.4%, 1/256 cases; p=0.0013, one-sided Fisher’s exact test; Fig. 2B; Table 2). In contrast, GAAGGA repeat expansions were not significantly enriched in ALS at any threshold tested (Table 3).

**Figure 2.**
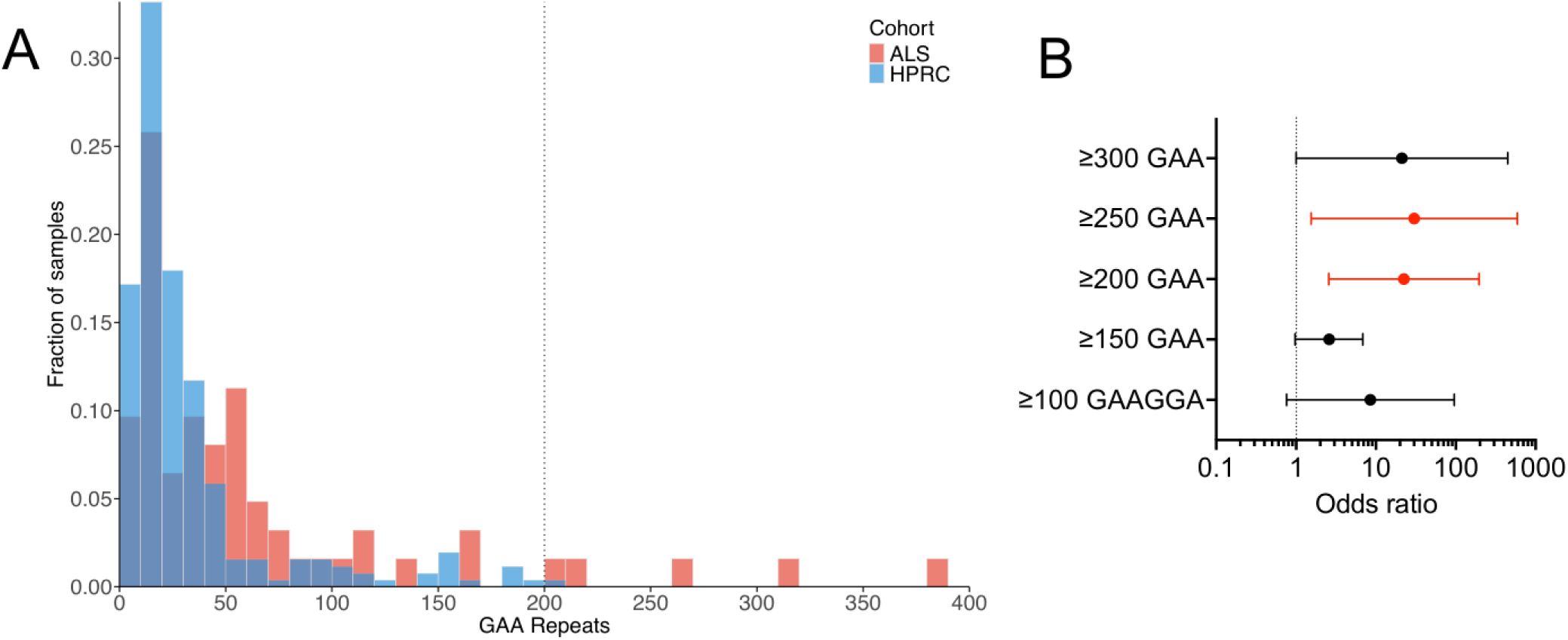
GAA repeat expansions in *FGF14* greater than 200 repeats are associated with ALS. (A) *FGF14* repeat length distribution of healthy control (blue; n=256; HPRC) and ALS cohorts (red; n=62). (B) Forest plot of the association between GAA and GAAGGA repeat sizes and ALS. Repeat sizes that are significantly associated with ALS are highlighted in red (p < 0.05, one-sided Fisher’s exact test). The odds ratio for ≥150 GAAGGA repeats is undefined due to the absence of ALS carriers in this repeat size category.

**Table 2.**
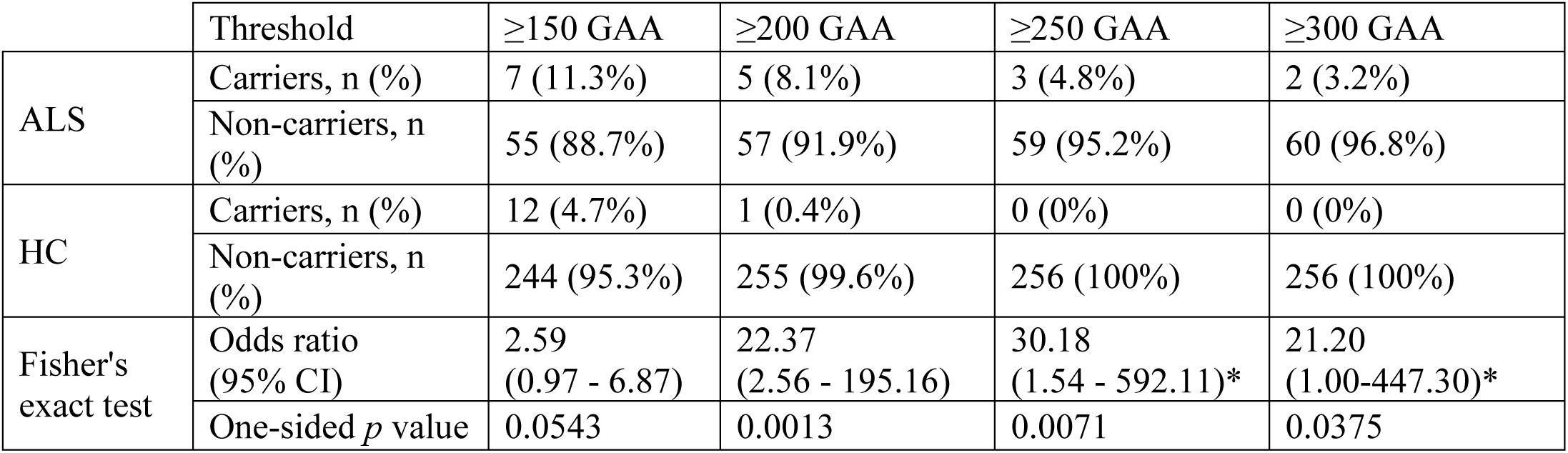
*FGF14* GAA repeat expansions greater than 200 units are significantly associated with ALS. Proportion testing and Fisher’s exact test were used to compare ALS versus healthy controls across thresholds binned by 50 repeats. *For ≥250 and ≥300 GAA thresholds, Haldane-Anscombe correction was applied to compute odds ratios and confidence intervals due to zero carriers in the control cohort.

**Table 3.**
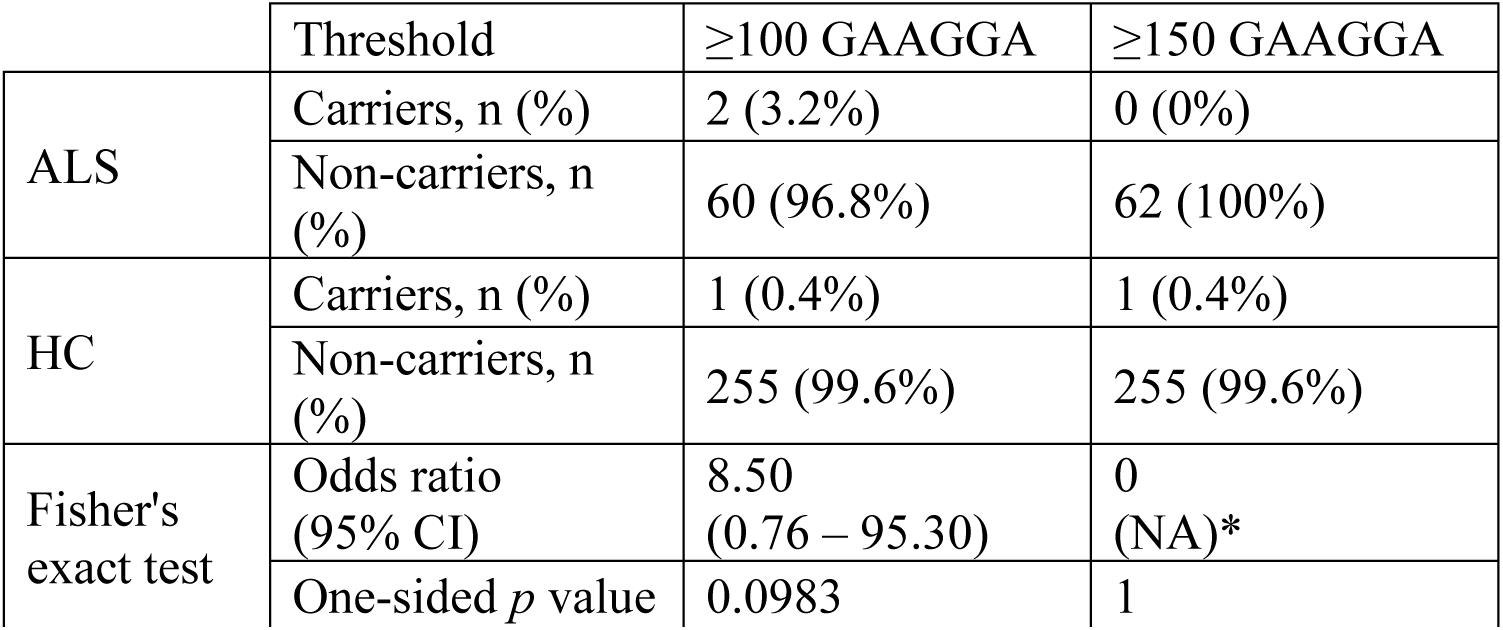
GAAGGA repeat expansions in *FGF14* are not significantly associated with ALS. Fisher’s exact test results are shown for thresholds binned by 50 repeats. *Confidence intervals were not defined for ≥150 GAAGGA due to the absence of ALS carriers in this repeat size category.

### Clinical assessment of ALS participants with *FGF14* repeat expansions

Clinical and demographic data were available for the three GAA expansion carriers with >250 repeats and two GAAGGA expansion carriers (Table 4). All cases were confirmed ALS diagnoses, with other disease processes excluded by neuroimaging and electrophysiological testing, and did not exhibit cerebellar symptoms nor downbeat nystagmus that are features of SCA27B. None of the cases carried a known ALS-associated genetic variant, nor did they have any family history of ALS. Two GAA expansion carriers with >250 repeats reside in USA and one participant resides in Australia; both GAAGGA expansion carriers reside in USA.

**Table 4.** Demographic and clinical features of *FGF14* GAA and GAAGGA expansion carriers.

| Carrier ID | GAA-1 | GAA-2 | GAA-3 | GAAGGA-1 | GAAGGA-2 |
| --- | --- | --- | --- | --- | --- |
| **LRS *FGF14* expansion** | 319, 12 GAA | 262, 12 GAA | 385, 43 GAA | 143, 0 GAAGGA | 144, 0 GAAGGA |
| **Age of onset** | 44 | 57 | 75 | 42 | 68 |
| **Gender** | M | F | F | M | F |
| **Race** | White | White | White | White | White |
| **El Escorial category** | - | Definite ALS | Possible ALS | Probable ALS | Probable ALS |
| **Family history of ALS** | No | No | No | No | No |
| **ALS-associated genetic variant carrier** | No | No | No | No | No |
| **Site of Onset** | Limb | Bulbar | Bulbar | Limb | Limb |
| **Duration, years** | 12.2 | 2.09 | 1.21 | 14.49 | 0.41 |
| **2024 ALSFRS-R** | - | 31 | 42 | 39 | 39 |
| **Progression rate (DALSFRS-R points/month)** | - | 0.56 | 0.67 | 0.56 | 1.33 |

### Repeat expansions of GAA in *FGF14* are predicted to form H-DNA triplex structures

To investigate the potential pathogenic mechanism associated with the expanded GAA repeat, we performed *in silico* analyses to predict the formation of non-B DNA structures that could impair transcription of *FGF14* and promote gene silencing, as described in other triplet repeat expansion disorders (25). QmRLFS-finder analysis predicted no evidence of R-loop formation in the expanded GAA regions of *FGF14*, suggesting that R-loop-mediated transcriptional repression is unlikely to be the primary pathogenic mechanism of this variant. We next employed the nBMST tool to search for additional non-B DNA structures. Expanded (GAA) alleles were predicted to contain multiple, dense clusters of long mirror repeats across the repeat tract, a sequence architecture known to facilitate the formation of H-DNA (triplex) structures (Fig. 3). Notably, the likelihood and stability of this predicted H-DNA structure increased proportionally with GAA repeat length. On the other hand, non-expanded (GAA) alleles and the mixed-motif allele (GAA)_20_(GAAGGA)_141_ exhibited short mirror-repeat predictions and lacked extended long-repeat clusters. Together, these analyses show that long, pure GAA expansions are uniquely predicted to accumulate extended mirror-repeat tracts, whereas short GAA or mixed-motif GAAGGA alleles do not, supporting the pathogenicity of long GAA repeat units.

**Figure 3.**
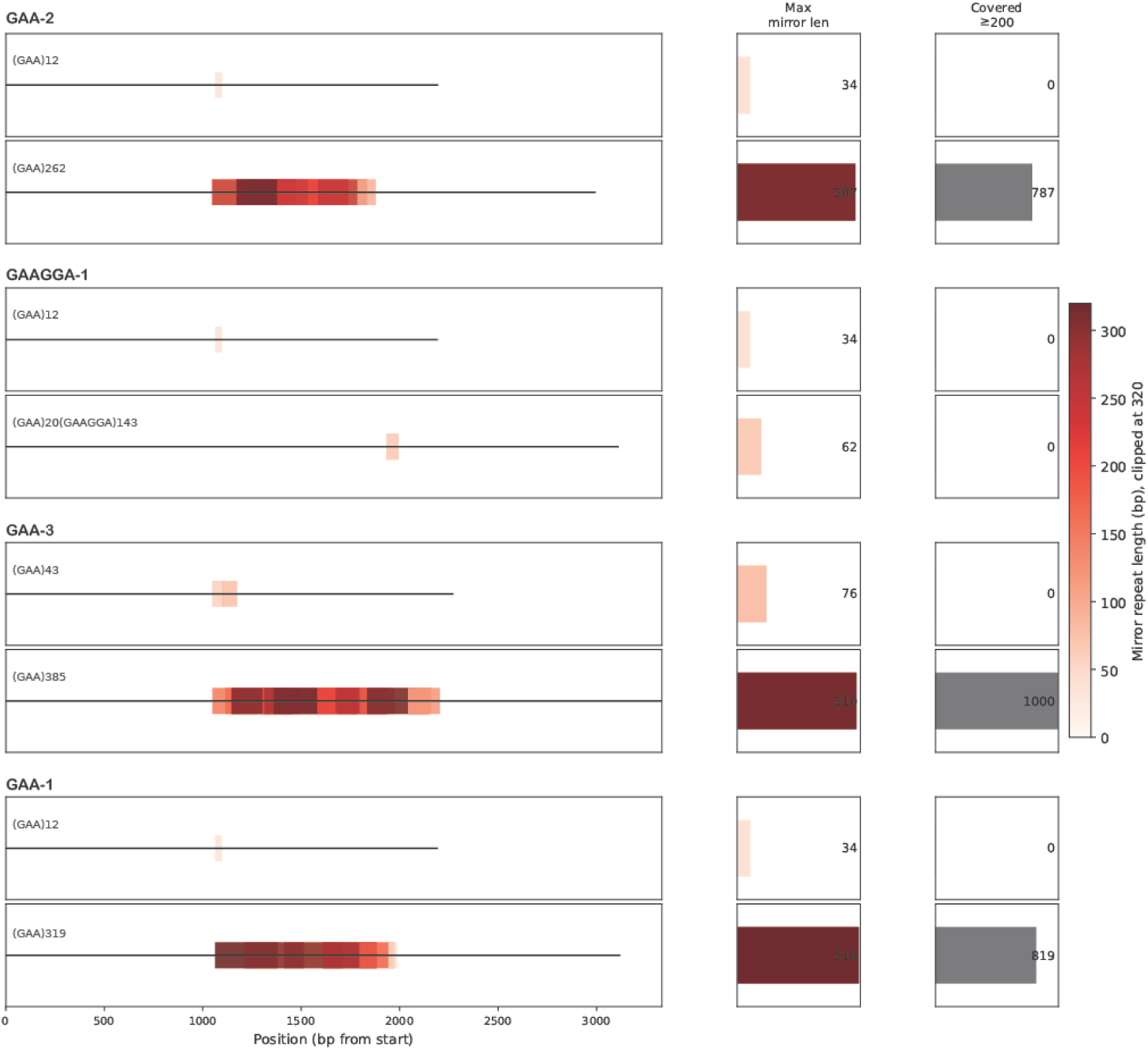
Repeat expansions of GAA, but not GAAGGA, in *FGF14* are predicted to form H-DNA (triplex-forming) structures. Predicted mirror-repeat regions identified by nBMST are shown as horizontal rectangles along each allele sequence (left panels). The black horizontal line represents the input sequence analysed for each allele (±1000 bp flanking the repeat expansion). Rectangle colour indicates mirror-repeat length (bp) on a continuous scale (0–320 bp; values above 320 are clipped). Right panels summarise mirror-repeat features per allele. Max mirror len denotes the longest predicted mirror repeat (bp) within the allele. Covered ≥200 reports whether long mirror repeats (≥200 bp) are present and, if so, the total number of base pairs spanned by the union of ≥200-bp mirror-repeat intervals (i.e., long-repeat coverage without double-counting overlaps).

### *FGF14* isoform expression in motor neurons

The *FGF14* gene is expressed as at least two major isoforms, 1A (canonical) and 1B (alternate). The structure of the two isoforms is identical except for their N-terminal coding regions, which are encoded by unique first exons (Supplementary Fig. 1). Notably, the GAA repeat expansion is located within an intronic region that is included in the longer isoform 1B transcript but is absent from the shorter isoform 1A transcript (14). To determine the biological relevance of these isoforms in the affected cell type, we analysed RNA-sequencing data from induced pluripotent stem cell-derived spinal motor neurons (iPSC-MNs) (22). To determine the relative expression levels of the *FGF14* isoforms, we compared the RNA-seq counts in the first exon of isoform 1A versus the first exon of isoform 1B. The results revealed an unexpected expression pattern, with almost all detectable *FGF14* gene expression associated with isoform 1B (Fig. 4A). On the other hand, the shorter, canonical isoform 1A, which does not span the GAA repeat tract, was expressed at very low levels in the iPSC-MN cell lines. This finding strongly suggests that isoform 1B carrying the pathogenic GAA repeat expansion is the biologically relevant transcript in spinal cord motor neurons. However, we detected no significant difference in *FGF14* mRNA levels in ALS versus control iPSC-MNs (Fig. 4B).

**Figure 4.**
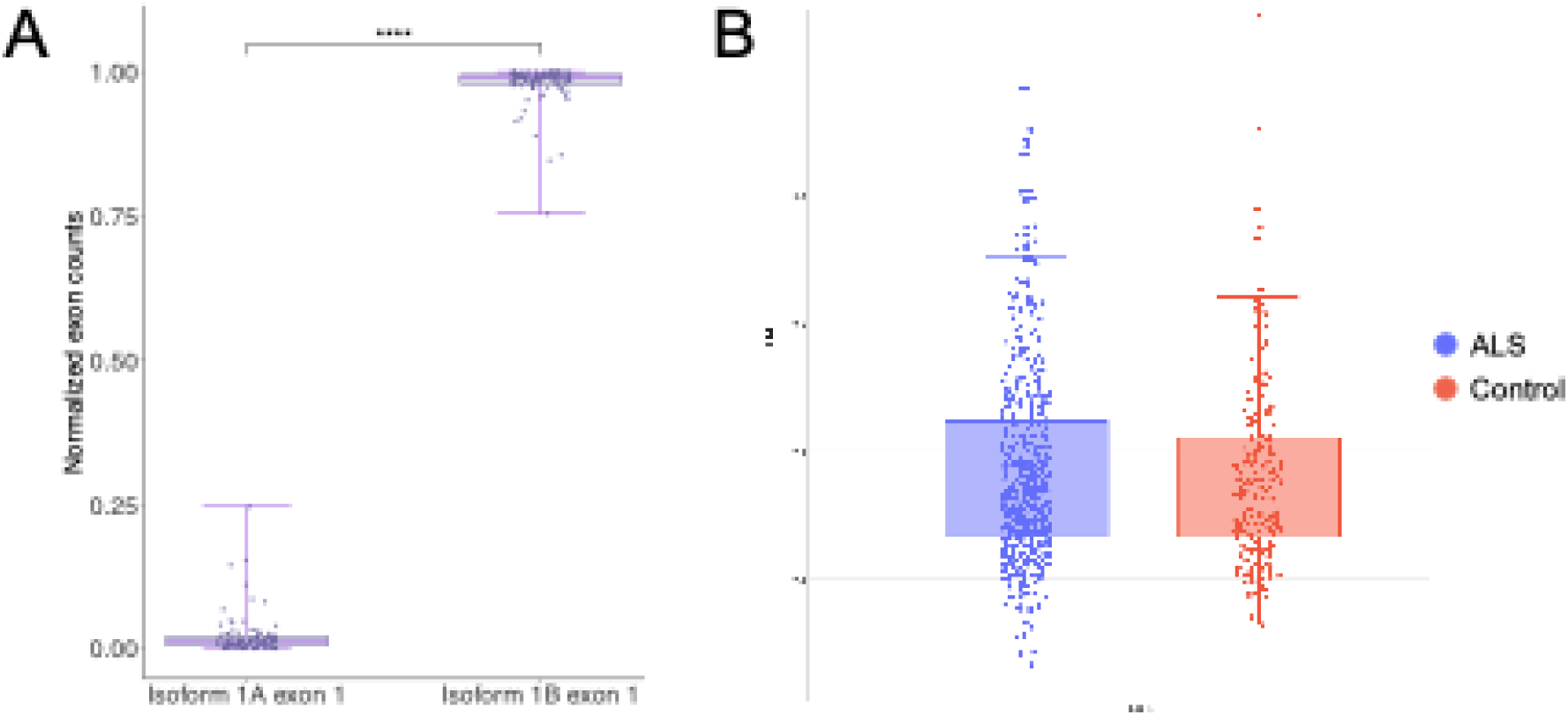
iPSC-derived motor neurons express the *FGF14* transcript that spans the GAA repeat tract. (A) Exon-level quantification of the first exon of *FGF14* isoform 1A versus the first exon of isoform 1B in iPSC-MNs (95 total: 28 controls, 67 ALS). The first exon of each transcript is unique to that isoform. ****, p < 0.0001 by paired Wilcoxon signed-rank test. (B) Analysis of *FGF14* mRNA levels in control (n=174) and ALS (n=458) iPSC-MNs using the AnswerALS Visualization Tool.

By contrast, gene expression analysis of bulk RNA-seq data from 101 human motor cortex samples demonstrated significantly increased *FGF14* expression in ALS compared with controls (log2 fold change 0.153; p=0.0035; Fig. 5).

**Figure 5.**
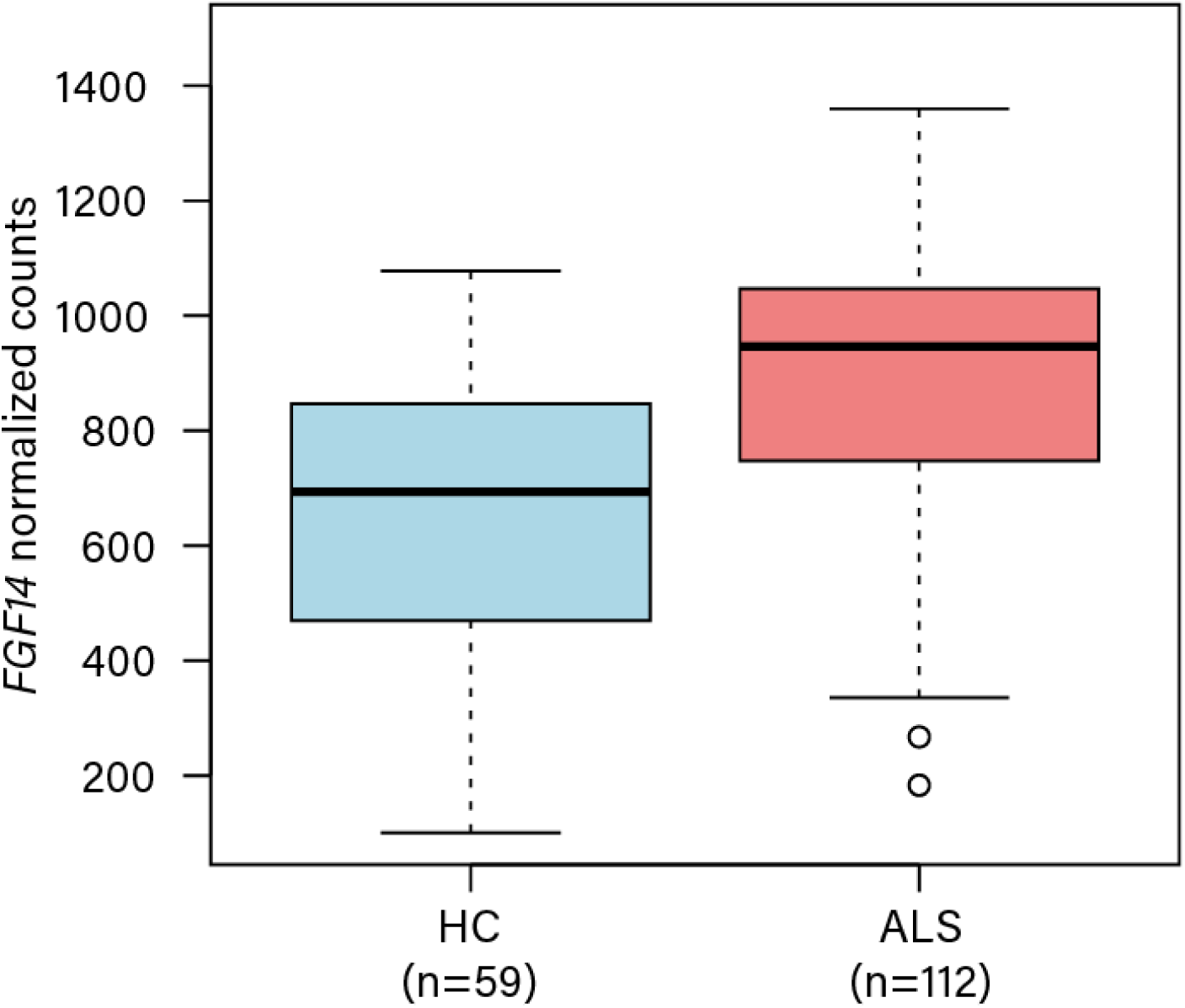
Increased *FGF14* expression in the motor cortex of participants with ALS. Bulk RNA-seq analysis of human motor cortex tissue (n=101; n=25 controls and n=76 ALS) shows increased *FGF14* expression in samples from individuals with ALS compared with controls (log2 fold change 0.153; p=0.0035). Gene expression levels are shown for individual samples; summary statistics and statistical testing details are provided in the Methods.

## Discussion

The identification of GAA and GAAGGA repeat expansions in the *FGF14* gene within an ALS cohort represents an important advance in understanding the genetic landscape of this neurodegenerative disorder. We identified pathogenic-range *FGF14* GAA ≥250 expansions, the established threshold for SCA27B(16, 24), in 3 of 62 ALS cases (4.8%) and their absence in healthy controls, which directly links *FGF14* to ALS. Furthermore, repeat-size distribution analysis revealed a significant enrichment of GAA expansions ≥200 repeats in ALS cases (8.1%) compared with controls (0.4%; p=0.0013). The low prevalence of GAA expansion (≥250 repeats) observed in controls is consistent with that of other studies that reported healthy control prevalences of 0.2% in Japan (n=455) (19, 20), 1.4% in a mixed control population in England, Canada, Germany, USA (n=1003) (19, 20), 0.6% in Australia (n=210) (14, 15), and 0.98% in Canada and Germany (n=408) (14, 15).

Our findings suggest a broader pathogenic spectrum for *FGF14* GAA repeats in ALS than previously recognised in SCA27B. While ≥250 repeats define the classical SCA27B phenotype, our data indicates that intermediate-to-large expansions (≥200 repeats) are significantly associated with ALS, implying that even shorter expansions may confer susceptibility or modify disease presentation in MND. This aligns with a broader trend in ALS genetics, in which intermediate and large repeat expansions at several loci contribute to ALS susceptibility in diverse populations. Recent work by Novy et al. (2024) in a Norwegian cohort underscores the importance of screening for repeat expansions in genes like *ATXN1*, *ATXN2*, and *HTT* highlighting that the genetic architecture of ALS is defined by a broad spectrum of repeat-related pleiotropy (26). Another recent study by Laß et al. reported complex mosaic interruptions of *FGF14* repeat expansions, in late-onset ataxia individuals, where motif length and sequence were associated with a protective effect in comparison to pure GAA repeats (27). Larger studies using long-read whole-genome sequencing for both people with ALS and healthy controls in ancestrally matched cohorts will be essential to refine the *FGF14* repeat size threshold that confers risk or pathogenicity in ALS.

GAAGGA repeat expansions are considered not pathogenic for SCA27B (14, 24) and did not appear to increase ALS risk in our cohort. Given the small cohort size of this study, no phenotypic associations with repeat expansion size or motif could be made. Motif-dependent variations in clinical presentation are not uncommon in repeat expansion disorders, as seen with repeat interruptions in *C9ORF72*(28) or the purity of repeats in other ataxias. Interruption of the SCA2 CAG repeat expansion with CAA repeats is associated with autosomal dominant parkinsonism rather than ataxia (29), while histidine interruptions in the polyglutamine tract of ATXN1 can prevent or mitigate SCA1 (30). Beyond motif variation, certain repeat expansions appear to have pleiotropic effects across the ataxia-ALS spectrum. Hirano et al. (2018) identified that noncoding expansions in the *ATXN8OS* gene, typically associated with spinocerebellar ataxia type 8 (SCA8), were present in 3% of Japanese sporadic ALS cases, with a notable clinical trend towards bulbar-predominant symptoms or neck weakness (31). The parallels between *FGF14* and *ATXN8OS* suggest that certain noncoding repeat expansions, while primarily linked to cerebellar phenotypes, may also possess a latent capacity to drive motor neuron degeneration in ALS, often with a preference for bulbar involvement. Larger-scale screening of *FGF14* repeat expansions in ALS populations should include collection of clinical and biomarker data to confirm whether distinct *FGF14* repeat expansion motifs indeed modify the clinical phenotype of disease.

Transcriptomic data revealed that isoform 1B is the predominant *FGF14* transcript in spinal cord motor neurons, while the canonical isoform 1A is nearly absent. This is a crucial finding because the repeat expansion is located within intron 1 of isoform 1B. The dominance of this isoform in spinal motor neurons, the disease-relevant cell type in ALS, strengthens the plausibility of *FGF14* as a causative gene in ALS. Interestingly, we observed increased *FGF14* expression in ALS human motor cortex tissue compared with controls, but not in iPSC-derived motor neurons. This discrepancy may reflect compensatory upregulation in the adult brain during the disease process or age-related changes that are not modelled by relatively immature iPSC-derived models. Other data also indicate elevated levels of FGF14 in ALS-blood (Human Protein Atlas; proteinatlas.org) (32). Interestingly, the levels of *FGF14* mRNA and protein are downregulated in both post-mortem cerebellar tissues and in iPSC-MNs from SCA27B patients compared with controls, suggesting that the intronic expansion causes loss-of-function as result of impaired *FGF14* transcription in SCA27B (14). To further investigate the biological consequences of carrying the *FGF14* repeat expansion, future RNA-seq studies should compare people with ALS and SCA27B who are carriers versus non-carriers.

Molecular analyses provided insights into the potential pathogenic mechanisms of these expansions. *In silico* modelling did not predict formation of R-loops, but instead strongly predicted the formation of H-DNA (triplex) structures in alleles with long GAA repeats. On the other hand, short GAA or mixed-motif GAAGGA alleles were not predicted to significantly impact the structural integrity of DNA. This aligns with previous work demonstrating that *FGF14* GAA repeats form unique secondary structures at the DNA and RNA level (24). H-DNA structures have been demonstrated in other repeat expansion diseases such as in *FXN* in Friedreich’s Ataxia, where they induce genomic instability, facilitate further repeat expansion, and interfere with transcriptional processes, resulting in loss of function (33, 34). The positive correlation between expansion length and the likelihood of H-DNA formation also suggests a mechanism for anticipation or somatic instability, common features in repeat expansion disorders (35).

Although our findings highlight a significant role for *FGF14* repeat expansions in ALS, several limitations warrant consideration. Firstly, the relatively small sample size of repeat expansion carriers underscores the need to replicate this finding in larger and more diverse ALS cohorts to establish the true prevalence and clinical relevance of *FGF14* repeat expansions in this disease. Secondly, while our genomic data support the presence of an unstable GAA repeat that can adopt non-B-DNA conformations, experimental validation of H-DNA formation and transcriptional attenuation in the *FGF14* locus is required to substantiate the proposed pathogenic mechanism. Finally, the precise molecular consequences of the *FGF14* repeat expansions in ALS, particularly their impact on FGF14 protein levels and its function, as well as downstream cellular pathways, remain to be elucidated. Addressing these gaps will be essential to define how *FGF14* repeat expansions contribute to ALS pathophysiology and for determining their potential utility as diagnostic markers or therapeutic targets.

In conclusion, our study demonstrates that *FGF14* repeat expansions extend into the MND spectrum, identifying them as a novel genetic risk factor for ALS. These findings highlight a shared genetic architecture between ALS and other neurodegenerative disorders like SCA27B. By identifying a broader pathogenic repeat spectrum, this work emphasises that the phenotypic boundary between ataxia and other motor neuron diseases is increasingly fluid, mediated by the complex interplay of repeat motifs and isoform-specific expression. Future research should focus on elucidating the specific functional consequences of these expansions on motor neuron physiology and whether targeting *FGF14* or its downstream effectors could offer a new therapeutic strategy for people with ALS.

## Acknowledgments

We thank people living with ALS for participating in this study and for donations which partially funded this work. Data used in the preparation of this article were obtained from the ANSWER ALS Data Portal (AALS-01184). For up-to-date information on the study, visit https://dataportal.answerals.org. AAC is an NIHR Senior Investigator (NIHR202421) and a Visiting Professor at the Perron Institute for Neurological and Translational Science, Australia. AAK and AI are Visiting Senior Fellows at the Perron Institute for Neurological and Translational Science, Australia. We are grateful to the London Neurodegenerative Diseases Brain Bank for providing the samples used to generate the King’s College London RNAseq dataset. The authors acknowledge use of the King’s Computational Research, Engineering and Technology Environment (CREATE) (https://create.kcl.ac.uk), which is delivered in partnership with the National Institute for Health and Care Research (NIHR) Biomedical Research Centres at South London and Maudsley and Guy’s and St. Thomas’ NHS Foundation Trusts and part-funded by capital equipment grants from the Maudsley Charity (award 980) and Guy’s and St. Thomas’ Charity (TR130505).

## Author contributions

SM and MS conceived and designed the study and wrote the manuscript with input from all authors. PKW analysed data and edited the manuscript. AT performed the genome processing and AnswerALS RNAseq data analysis. AY performed the R-loop and H-DNA analyses. ED performed the Human Pangenome Reference Consortium data analysis. AAK performed the King’s College London RNAseq data analysis. TW performed genome processing. PAA assisted with collaborations. NE-O supported the genome processing. MF analysed data and edited the manuscript. MBH assisted with study design and edited the manuscript. THP and RB acquired participant data. All authors reviewed and approved the final version.

## Web Resources

ANSWER ALS Data Portal (AALS-01184), https://dataportal.answerals.org

Human Pangenome Reference Consortium, https://humanpangenome.org/

GTEx Portal, https://www.gtexportal.org/

QmRLFS-finder, http://r-loop.org/?pg=qmrlfs-finder

## Data and code availability

Sequencing data have not been deposited in a public repository due to restrictions associated with participant consent but are available from the corresponding author on reasonable request.

