## Supplementary Table 1, Supplementary Figure 1, Supplementary Figure 2 for "Enrichment of Repeat Expansions in *FGF14* Associated with Amyotrophic Lateral Sclerosis"

| **Group** | **n** | **Sex (M:F)** | **Age at death, years (SD)** | **PMD, h (SD)** | **RIN (SD)** | ***C9ORF72* HREM, n (%)** |
| --- | --- | --- | --- | --- | --- | --- |
| ALS | 76 | 41:35 | 68 (8) | 26 (13) | 6.3 (1.3) | 5 (6.6%) |
| Control | 25 | 13:12 | 66 (9) | 36 (22) | 5.3 (1.7) | 0 (0%) |

**Supplementary Table 1: Donor characteristics for bulk RNA-seq analysis of post-mortem motor cortex from ALS cases and controls**. Values are shown as mean (SD) unless otherwise stated. PMD denotes post-mortem delay. RIN indicates RNA integrity number. *C9ORF72* HREM refers to the number of patients positive for a pathogenic hexanucleotide repeat expansion in *C9ORF72*.

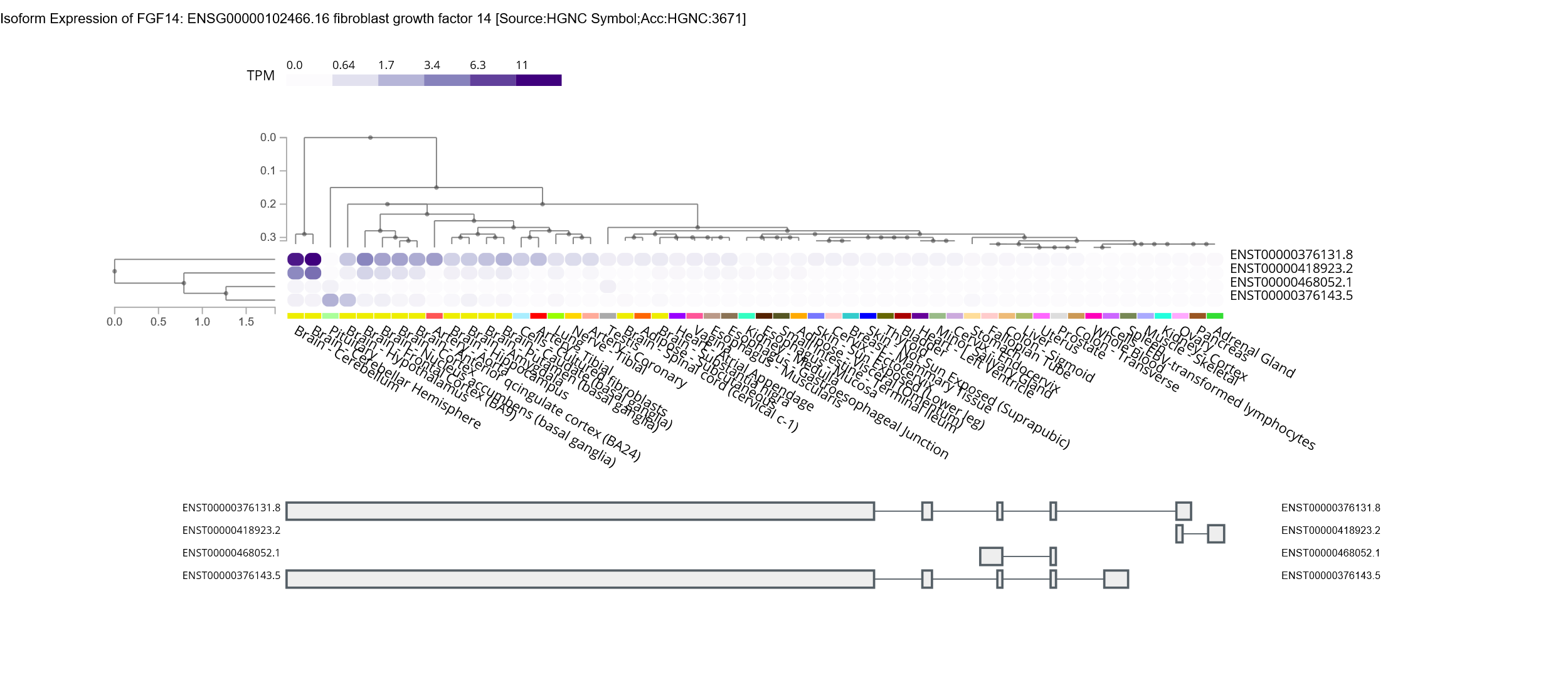
**Supplementary Figure 1. Isoform 1B is the dominant *FGF14* transcript expressed in the brain and spinal cord.** Expression of *FGF14* isoforms in the GTEx dataset. Isoform 1A is ENST00000376143.5 and Isoform 1B is ENST00000376131.8. *FGF14* occurs on the reverse strand of DNA and therefore exon 1 of each transcript is represented by the exon furthest on the right.

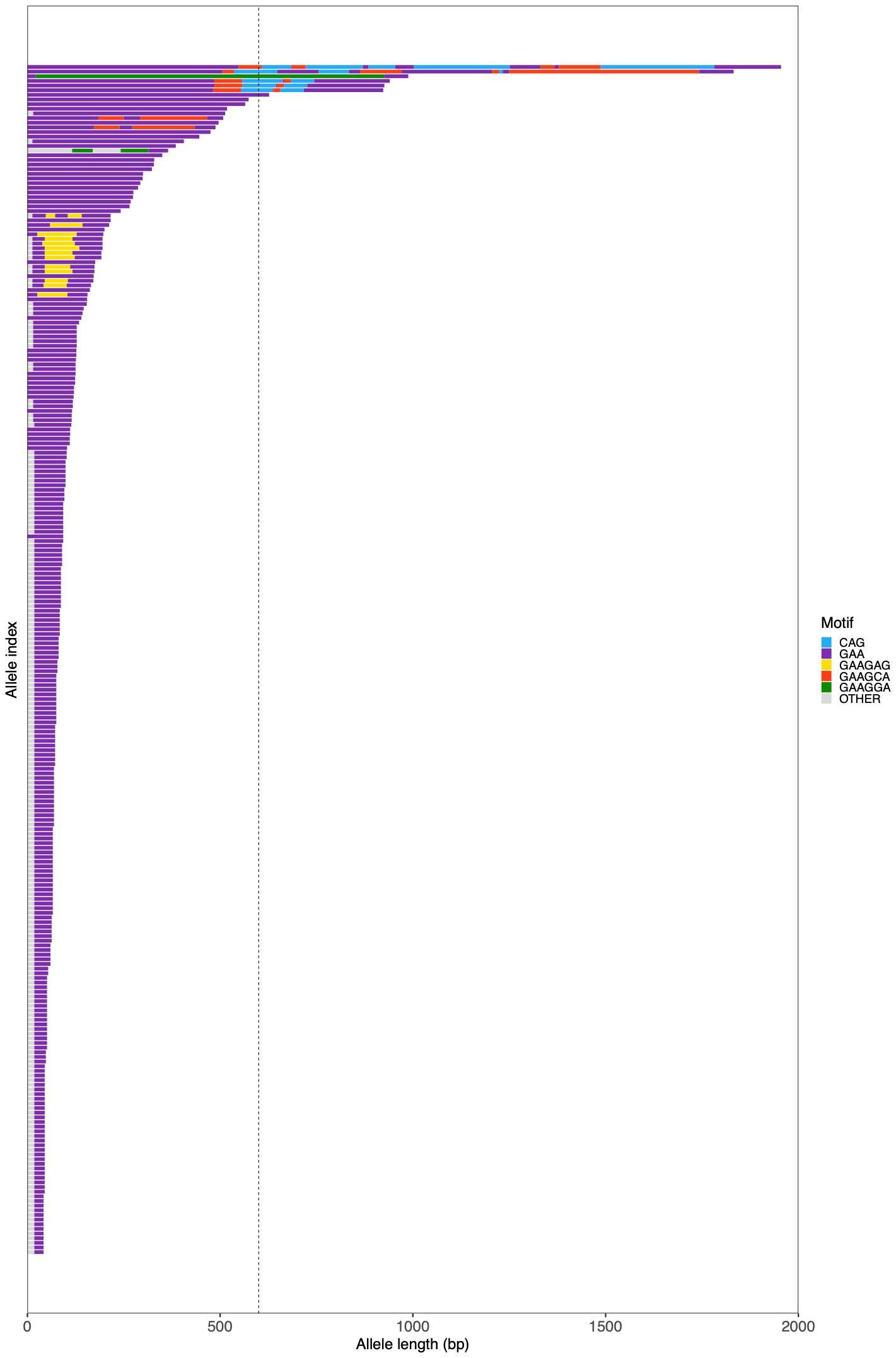

**Supplementary Figure 2. Characterization of the size and motif distribution of the *FGF14* repeat expansion in a cohort of 256 healthy individuals (Human Pangenome Reference Consortium).** The dashed line indicates an allele length of 600 bp (200 GAA repeats).
